# Proteogenomic analysis provides insights into the effect of physical activity on cancers

**DOI:** 10.64898/2026.11.25.26368973

**Authors:** Andrew J. Song, Chandrima Chatterjee, Steven Moore, Diptavo Dutta

## Abstract

**Background:** Increased physical activity (PA) has been associated with decreased risk of several cancers in epidemiologic studies, but the intermediate molecular mechanisms remains poorly characterized.

**Methods:** Leveraging genome-wide association study (GWAS) summary statistics for 22 self-reported and accelerometer-based PA measures, plasma levels of 2,940 proteins (N = 34,557), and multiple site-specific cancers, we performed a two-stage proteome-wide Mendelian randomization (MR) analysis. In Stage 1, we estimated the effects of each PA measure on plasma protein levels and in Stage 2, we evaluated the effects of PA-associated proteins on cancer risk.

**Results:** We identified 687 PA–protein associations involving 503 unique proteins at FDR<0.05. Across 80 PA-responsive proteins curated from intervention-based studies, we nominally replicated 83% using our MR approach. Approximately 40% of PA-associated proteins were differentially expressed in response to exercise in transcriptomic datasets primarily localizing to muscle tissues, and in immune cell populations. In stage 2, 213 (of 503) PA-related proteins demonstrated significant effects on at least one cancer (FDR<0.05) highlighting them as proteomic intermediates of the effect of PA on cancer. For example, for prostate cancer (PCa), we observed that reductions in sedentary time and increases in light-intensity activity were associated with higher MSMB and lower TNF levels, which in turn reduced PCa risk.

**Conclusions:** This study applies a two-stage Mendelian randomization framework to link PA, circulating proteins, and cancer risk, enhancing biological interpretation of epidemiologic PA-cancer associations and nominating a focused set of PA-associated proteins as potential targets for further investigation.

## Introduction

Cancer prevalence and mortality in the United States presents a major health burden, with over two million new cancer diagnoses and 611,720 deaths in 2024. Epidemiological and genetic studies, including genome-wide association studies (GWAS), have identified several risk factors and genetic susceptibility loci associated with different cancers^1,2^. Leveraging the findings from such studies for prevention, remains a central goal of cancer epidemiology. Among such factors, physical activity (PA) has shown consistent evidence for reduced risk of several disease^3–5^, including reduced risk of cancers^6,7^. PA is a multifaceted behavioral exposure rather than a single, uniform exposure. It encompasses the overall volume, intensity, and temporal distribution of activity, as well as sedentary behavior, which carries distinct behavioral, metabolic, and circadian correlates. These dimensions may relate to cancer risk through overlapping as well as distinct biological processes, and their effects might not be concordant. However, there is a limited understanding of the biological mechanisms through which different facets of PA influence carcinogenesis. In this respect, delineating the intermediate molecular processes that connect PA to cancer is crucial to deepen etiologic understanding, prioritize pathways for intervention, and inform prevention strategies. Characterizing the molecular correlates of PA therefore requires attention to these different facets rather than treating PA as a monolithic exposure.

A key consideration in delineating these mechanisms is the nature of the PA exposure that is relevant to cancer. Cancer is a long-latency disease, developing over years or decades through the gradual accumulation of inflammatory, metabolic, immune, and proliferative changes. Consequently, the PA-related exposure most relevant to carcinogenesis is not any single episode of exercise, but the sustained, habitual pattern of activity and sedentary behavior maintained over the life course. This distinction is critical and notable for etiologic and mechanistic understanding. Much of the existing evidence of the effects of PA on cancers, derives from candidate biomarker and exercise intervention studies, which characterize acute or short-term responses to prescribed, often higher-intensity exercise, and are frequently limited in size or to selected biomarker panels. While such designs are well suited to defining the immediate physiology of exercise, the transient responses they capture may not reflect the durable biological differences that distinguish habitually active from habitually inactive individuals. An approach that captures the molecular signatures of long-term, habitual PA is therefore needed to interpret its effects on cancer and to understand the intermediate molecular phenotypes that cascade the effects.

With the advent of high throughput technologies, circulating plasma proteins have gained prominence as molecular intermediates for integrative analyses of cancers^8–10^. Circulating proteins participate in fundamental processes implicated in cancer development such as inflammation, immune surveillance, metabolism, and hormonal signaling^11^. With the current technology, they are measurable at scale, can capture systemic responses across multiple tissues and cell types, and include many druggable targets, making them directly relevant for translational applications. Population-based proteogenomic studies have demonstrated that genetic regulation of plasma proteins can inform disease etiology and identify potential therapeutic targets across cardiometabolic, neurologic, and other complex diseases^8–10,12,13^. Extending such approaches to cancer-relevant risk factors offers a path to connect exposures to pathways and cancer outcomes within a unified, genetically anchored framework^14,15^.

Mendelian randomization (MR) provides a genetically anchored approach to characterize the effects of PA on the proteome as an intermediate^16–18^. MR utilizes germline genetic variants as instrumental variables (or proxies) for an exposure and tests their association with an outcome, and compared with traditional observational studies, has the conceptual advantage of being more robust to confounding and reverse causation^19^. Importantly, because germline variants are fixed from conception, genetic instruments for PA proxy a lifelong propensity toward higher activity and lower sedentary behavior, rather than a short-term change in activity in adulthood. In this way, MR indexes the sustained, habitual exposure that is most relevant to a long-latency outcome such as cancer, and can complement intervention studies, which characterize acute exercise responses. We thus interpret MR estimates of the effect of PA on circulating plasma proteome as reflecting durable, habitual differences in activity and sedentary behavior.

Here we conducted a two-stage MR analysis to identify the proteomic intermediates of the effects of PA on different cancers (**Figure 1**). In stage 1, we estimated the effects of 22 accelerometer-derived and self-reported PA measures on plasma levels of 2,940 proteins. In stage 2, we evaluated the effects of PA-associated proteins on risk of multiple site-specific cancers and subtypes, thereby prioritizing candidate proteomic intermediates linking PA to cancer. We further benchmarked the PA-protein associations against proteins previously reported to respond to exercise in intervention studies and performed downstream in silico analyses to contextualize the prioritized proteins across pathways, cell types, and tumors by integrating external data. Collectively, this study provides a genetically anchored map of proteins most likely to reflect sustained, habitual PA and a focused set of candidate proteomic intermediates for mechanistic follow-up and further laboratory investigation.

**Figure 1:**
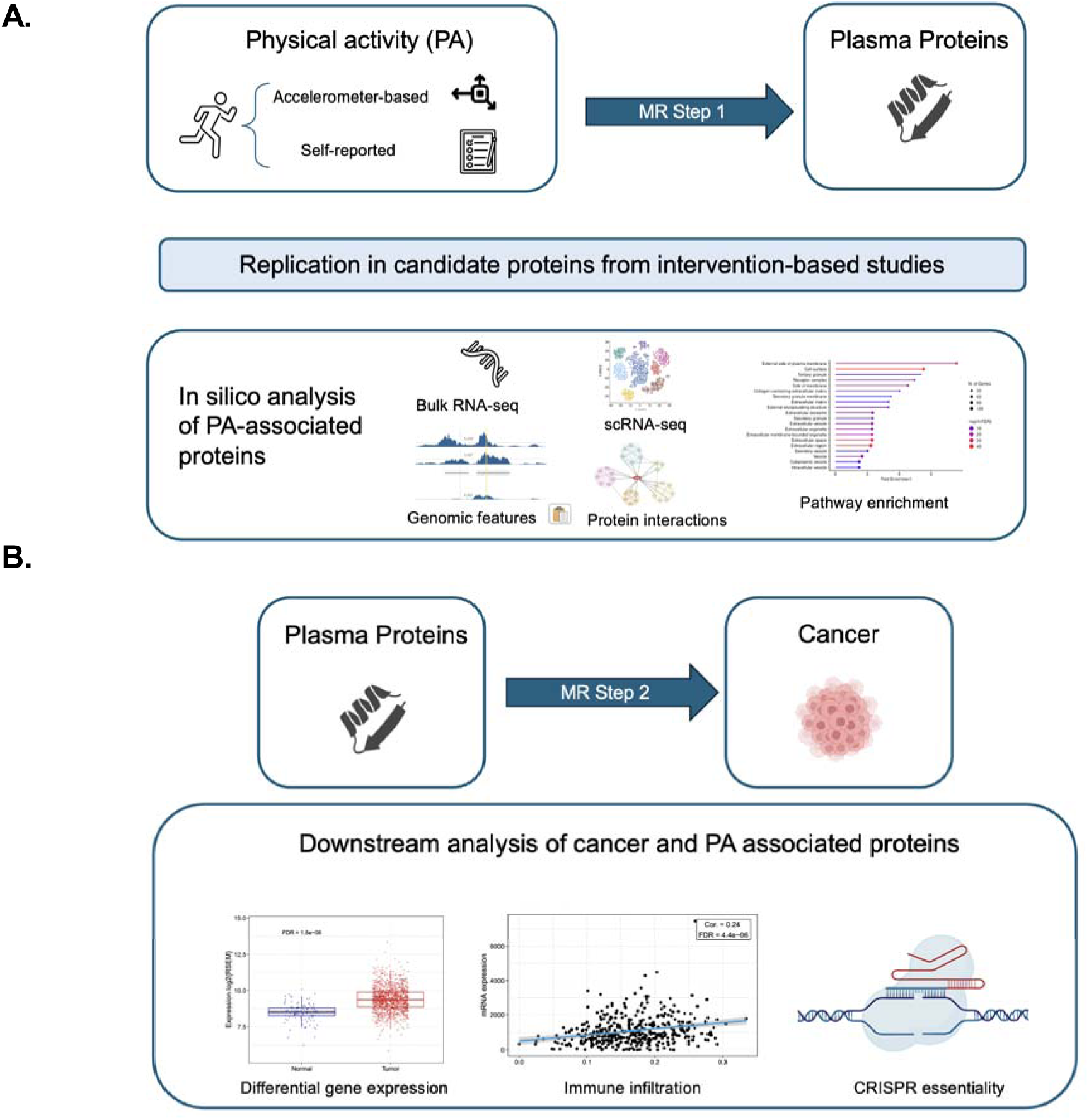
Overall schema of the analysis to identify PA-related proteins associated with cancers. (A) MR to identify effect of accelerometer-based and self-reported PA on plasma proteins along with further replication of intervention studies of PA and ultiple in silico analysis to establish the relevance of identified PA-related proteins. (B) MR to identify effect of PA-associated proteins on cancers along with downstream analysis to evaluate the effect of the identified proteins on respective cancers using tumor profiling.

## Methods

For our primary analysis, we performed a two-stage mendelian randomization (TSMR) analysis using publicly available GWAS summary statistics, stage 1: effect of PA on plasma proteome and stage 2: effect of PA-related proteins on different cancers. We first describe the details of MR analyses, which remained broadly the same across the two stages. All contributing studies obtained ethics approval and informed consent from participants as reported in the original publications. Our analyses used de-identified, publicly available summary statistics and did not require additional institutional review.

### Identifying genetic instruments

We restricted MR analyses to bi-allelic variants, common (minor allele frequency [MAF] > 1%) in GWAS for both the exposures and the outcomes. Further, allele frequencies for genetic variants were compared to the 1000 Genomes EUR reference panel (N = 498)^20^ and variants with mismatched alleles were removed from subsequent analyses. For each PA, genetic instruments were identified through linkage disequilibrium (LD)-clumping resulting in independent SNPs associated with the exposures (p-value < 5×10^−6^ for PA and < 5×10^−8^ for proteins). Correlated SNPs were excluded based on measures of LD (R^2^ > 0.1) and only SNPs with the most significant p-value were retained as genetic instruments. Subsequently, the instrumental variables for each PA were analyzed to estimate the effect of PA on protein using generalized summary statistics based Mendelian randomization (GSMR; see below).

<u>Removing pleiotropic SNPs:</u> We employed Heterogeneity in Dependent Instruments (HEIDI)^21^ to identify and address potential pleiotropy in genetic instruments. Briefly, the HEIDI outlier removal method tests for heterogeneity in the causal estimates derived from individual genetic instruments by measuring the inconsistency in the estimated effects across variants. To retain statistical power to detect heterogeneity, we used the default threshold of 0.01 meaning that even in the absence of pleiotropic outliers, only ∼1% of the genetic instruments will be removed by chance, which is unlikely to result in a substantial decrease in power of the subsequent MR analysis.

### Mendelian Randomization

We performed MR using GSMR^21^ which can efficiently handle summary-level data from GWAS and uses the inverse variance weighted (IVW) method to estimate the effect of a risk factor on the outcome. Using GWAS summary statistics from both the risk factor and outcome, GSMR identifies genetic instruments via clumping, removes potentially pleiotropic SNPs (via HEIDI) and uses the final list of genetic instruments to perform MR. GSMR further accounts for mild LD (r^2^ < 0.1) between the variants beyond LD clumping, estimated from an appropriate ancestry-matched reference panel. MR inference relies on instrumental variable assumptions: (1) genetic instruments are associated with the exposure (PA traits or protein levels); (2) instruments are independent of confounders of the association between exposure and outcomes; and (3) instruments influence the outcome only through the exposure (no horizontal pleiotropy). To evaluate the robustness of our analyses to violations of these assumptions, we calculated the F-statistic for instrument strength and also compared the significant results with those obtained from MR-Egger. For two-sample MR, we additionally assume comparable ancestry and alignment of genetic effects between exposure and outcome GWAS and correct for bias from sample overlap. All MR estimates are reported per 1-SD increase in genetically predicted exposure (PA or plasma proteins), with cancer outcomes expressed as log odds ratios.

#### TSMR Stage 1: Effect of PA on plasma proteome

We performed summary-based MR using different measures of PA as the exposure and circulating plasma protein levels as outcomes (**Figure 1A**). We collected a list of 22 different measures of PA, measured using accelerometer^22^ and questionnaires^23^ (**Table S1**) from two studies with available GWAS summary statistics. Given that effects of genetic variants on PA can be confounded by BMI and other anthropometric factors, we further conditioned on 4 anthropometric traits (BMI, WHR, body fat percentage and whole body fat mass)^24,25^ to obtain estimates of direct effects of SNPs on PA. For GWAS of plasma protein levels, we used publicly available GWAS summary statistics reported by UK Biobank Pharma Proteomics Project (UKB-PPP), across 2,940 proteins measured using Olink^8^. We utilize the reported GWAS in 34,557 individuals of European ancestry from the discovery cohort. The details of this proteomic GWAS have been reported previously^8^. For each PA, we defined a false discovery rate threshold of 5% to identify statistically significant PA-protein associations.

#### TSMR Stage 2: Effect of PA-related proteins on cancers

We performed summary-based MR using PA-related plasma protein levels as exposures and 8 different cancers, with publicly available summary statistics, as outcomes (**Figure 1B**). We collected data from five different studies with available GWAS summary statistics. Sample sizes ranged from 14,803 to 156,319 cases and from 27,065 to 944,762 total individuals of predominantly European ancestry. We further analyzed available subtypes of lung cancer, which included squamous cell carcinoma, adenocarcinoma, and lung cancer among smokers, and renal cell carcinoma, which included clear cell and papillary (**Table S1**). Similar as above, for each cancer, we defined a false discovery rate threshold of 5% to identify statistically significant protein-cancer associations.

#### Correction for sample overlap

MR estimates can be subject to potential biases due to overlap between exposure and outcome samples. In our study, GWAS for both physical activity and proteins included individuals from the UK Biobank. To account for this sample overlap, we used MRlap^26^, a software package, to correct for bias in PA and protein associations. For associations with significant difference in effect size before and after correction (p_diff_< 5×10^−3^), we used the bias corrected estimates from MRlap.

#### Benchmarking against exercise-intervention proteomics

Because genetic instruments for PA proxy durable, habitual differences in activity rather than acute exercise, we benchmarked Stage 1 against proteins reported to respond to exercise in intervention studies to test whether genetically proxied habitual PA recovers established exercise-responsive biology. We curated 195 genes and proteins previously reported to respond to exercise-related interventions or exercise-relevant processes, including heat-shock, autophagy, cardiorespiratory, and neurocognitive proteins from prior proteomic studies^27–31^. Of these, 84 genes or proteins had available UKB-PPP GWAS data and were carried into MR. We interpret this analysis as a benchmark rather than a direct replication: intervention studies typically measure acute, short-term, often higher-intensity exercise responses, whereas our genetic instruments proxy sustained, habitual activity and sedentary behavior, so partial rather than complete overlap is expected. For proteins associated with multiple PA traits, we summarized the strongest association.

## Results

### TSMR Stage 1: Effect of PA on plasma protein levels

#### Genetically proxied habitual PA recovers exercise-responsive proteins

We first assessed whether genetically proxied habitual PA recovers proteins known to respond to exercise, benchmarking the validity of PA instruments before proteome-wide analysis. Of 195 curated exercise-responsive proteins, 84 had available UKB-PPP genetic instruments. Their strongest associations yielded 74 significant PA-protein associations (p<0.05) involving 72 unique proteins, and 21 associations at p<1×10^−3^ (**Table S2-3**). This significantly higher than expected by chance. Even under the conservative assumption that all 22 PA measures are independent, fewer than 57 of the 84 proteins would be expected to show a nominal association, and the observed enrichment remains significant (p=1.3×10⁻☐). Of the 74 significant associations 43 involved accelerometer-derived activity measures, 27 involved self-reported leisure screen/sedentary time and 4 involved moderate-to-vigorous PA, indicating that genetically proxied habitual activity and sedentary behavior, can recover experimentally identified exercise-responsive proteins as well. Several associations were directionally relevant with the known biology of sustained activity. For example, leptin (LEP), an adipokine that declines with habitual activity^27,32^, was inversely associated with device-measured activity, including total log acceleration (TLAC_mean; β=−0.12, p=9×10⁻☐), light intensity activity (LIPA_Mins_mean; β=−0.17, p=2×10⁻☐) and evening activity between 6-8PM (TLAC_10_mean; β=−0.21, p=2×10⁻☐) with the direction of effects being consistent with PA-related improvements in energy balance and cardio-respiratory fitness. This benchmarking demonstrates that MR can realistically recapitulate experimentally observed proteomic effects of PA, supporting its utility as an *in silico* framework for testing potentially causal hypotheses in molecular exercise biology.

#### Proteome-wide effect of PA

Encouraged by the high level of corroboration of intervention-based associations by MR results, we next expanded our investigation to identify the effects of PA measures on a proteomic scale. Across PA traits, the median F-statistic for instrument strength was > 22 indicating low bias due to weaker instruments. We identified 687 unique associations between PA and proteins (**Figure 2A-B**), comprising 503 unique proteins (FDR < 0.05). Analysis using MR-Egger produced similar estimates for significant associations, with the correlation between estimated effects being 83.1% (p< 2×10^−16^) and only 24 (4.7%) exhibiting nominal evidence (p-value of intercept < 0.05) of pleiotropy. Approximately half of the proteins (50.2%; n = 253/503) were significantly associated with at least one accelerometer-based PA measure (mean 49 proteins per measure), and 59.2% (298/503) with self-reported leisure screen/sedentary time (LST_mean), whereas self-reported moderate-to-vigorous activity (MVPA_mean) showed no FDR-significant associations (**Table S2**). Total log acceleration (TLAC_mean), an index of overall activity volume, had the most associations (176 proteins at FDR < 0.05) the strongest association being with ITGAV (β=0.23, p=3.4×10^−11^). Among the self-reported measures of PA, the most significant association was of LST_mean with NOTCH3 (β=-0.15, p=8.2×10^−12^).

**Figure 2:**
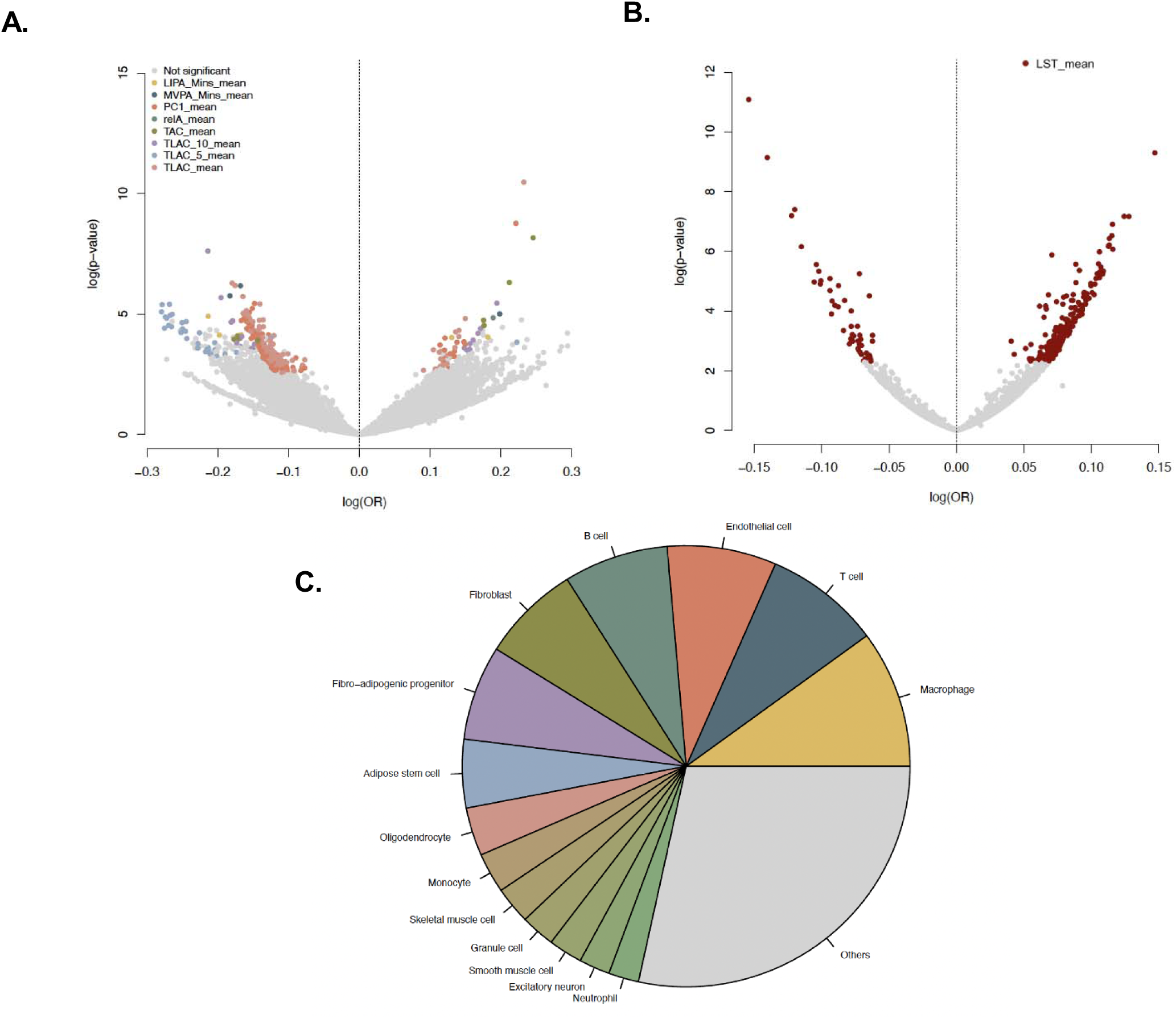
Effect of PA on plasma protein levels. Volcano plots showing the effect and p-value for associations between PA and proteins for (A) accelerometer-based PA measures (B) self-reported PA measure. Significant associations (FDR < 0.05) are colo ed by the respective PA measures. (C) Pie chart showing the most prominent cell types where the PA-associate proteins are differentially expressed.

Sedentary behavior was not simply the inverse of activity. Of 45 proteins associated with both self-reported sedentary behavior and at least one accelerometer-based PA measures, some tracked coherently across modalities. For example, ITGAV was inversely associated with sedentary time (LST_mean; β=−0.08, p=6.2×10⁻☐) and positively with activity volume. Similarly, MYOM3, a muscle-associated protein^33^, exhibited the lower association with sedentary time (β=−0.12, p=6.4×10⁻☐) and higher with evening activity (TLAC_10_mean; β=0.17, p=4.1×10⁻☐), supporting consistent PA-related effects across measurement modalities. There were 138 proteins associated with multiple PA measures, including previously discussed proteins like LEP which also exhibited response to PA in interventional studies, and several members of the tumor necrosis family (TNF, TNFRSF11A), growth factor (VEGFB) and multiple immunoregulatory proteins (CD22, CD72, C2). This indicates that distinct facets of habitual activity converge on a shared set of inflammatory, vascular, and immune proteins while retaining facet-specific signals..

#### Sensitivity to sample overlap

Because the PA and protein GWAS both drew on UK Biobank, we assessed bias from sample overlap using MRlap^26,34^. We found minimal evidence of substantial bias due to sample overlap, with MR estimates before and after correction for sample overlap being largely similar (r_pearson_ = 0.64). Of the 687 total PA-protein associations, 17.8% had nominal differences (p_diff_ < 0.05) between the MR estimates before and after correction by MRlap, and with a low proportion (5.6%) of PA-protein associations having stronger differences (p_diff_ < 0.005), that merited correction by MRlap (**Supplementary Figure**, **Table S4**).

#### In silico analysis in transcriptomic and pathway data

We contextualized the 503 proteins using ExerGeneDB^35^ a database of genes differentially regulated following exercise, PA and related exposures, across several laboratory experiments in humans and mice. Among the 503 PA-related proteins, nearly half (48.1%)corresponded to genes differentially expressed after exercise in at least one human bulk RNA-seq experiment, predominantly in muscle (97.5%) and colorectal mucosa (2.5%; (**Table S5**). In mouse single-cell data, 88% were differentially expressed in at least one cell type, most often macrophages (10%), T cells (8.4%), endothelial cells (7.9%), B cells (7.6%), and fibroblasts (7.2%; **Figure 2C**), pointing to immunoregulatory as well as metabolic processes. The 503 PA-related proteins showed significant protein–protein interaction enrichment (p<1.0 x 10^−16^; **Supplementary Methods**) and were overrepresented in several known pathways related to exercise. This included the Phosphatidylinositol 3-kinase (PI3K)/protein kinase B (Akt) signaling pathway (P_adj_=1.5×10^−11^), MAPK signaling (P_adj_=1.1×10^−06^), NF-kappa B signaling pathway (P_adj_=7.6×10^−6^) along with neuroimmune pathways such as neurogenesis (P_adj_=2.2×10^−07^), neuron differentiation (P_adj_=7.2×10-06), regulation of neuroinflammatory process (P_adj_=5.0 x 10^−5^) among others. In particular, pathways related to immune response were also highly enriched, including positive inflammatory response (P_adj_=2.1×10^−11^), T cell activation (P_adj_=8.3 x 10^−10^), B cell activation(P_adj_=2.4×10^−6^), among others.

### TSMR Stage 2. Effect of PA-associated proteins on cancer

We next tested whether the 503 proteins associated with habitual PA and sedentary behavior were associated with cancer risk, using summary-based MR with these proteins as exposures and 8 cancers and subtypes as outcomes. Of the 503 proteins, 213 (42.3%) had a significant effect on at least one cancer (FDR<0.05; **Figure 3A**). Because these proteins are supported in both stages, associated with habitual PA in Stage 1 and with cancer risk in Stage 2, we refer to them as candidate proteomic intermediates linking genetically proxied habitual PA to cancer, while noting we do not directly test formal mediation. The number of associated proteins varied across cancers, averaging about 27 per cancer and largely tracking the size of the corresponding GWAS: prostate cancer had the most (n=94), followed by breast (n=87) and colorectal cancer (n=56; **Figure 3B**).

**Figure 3:**
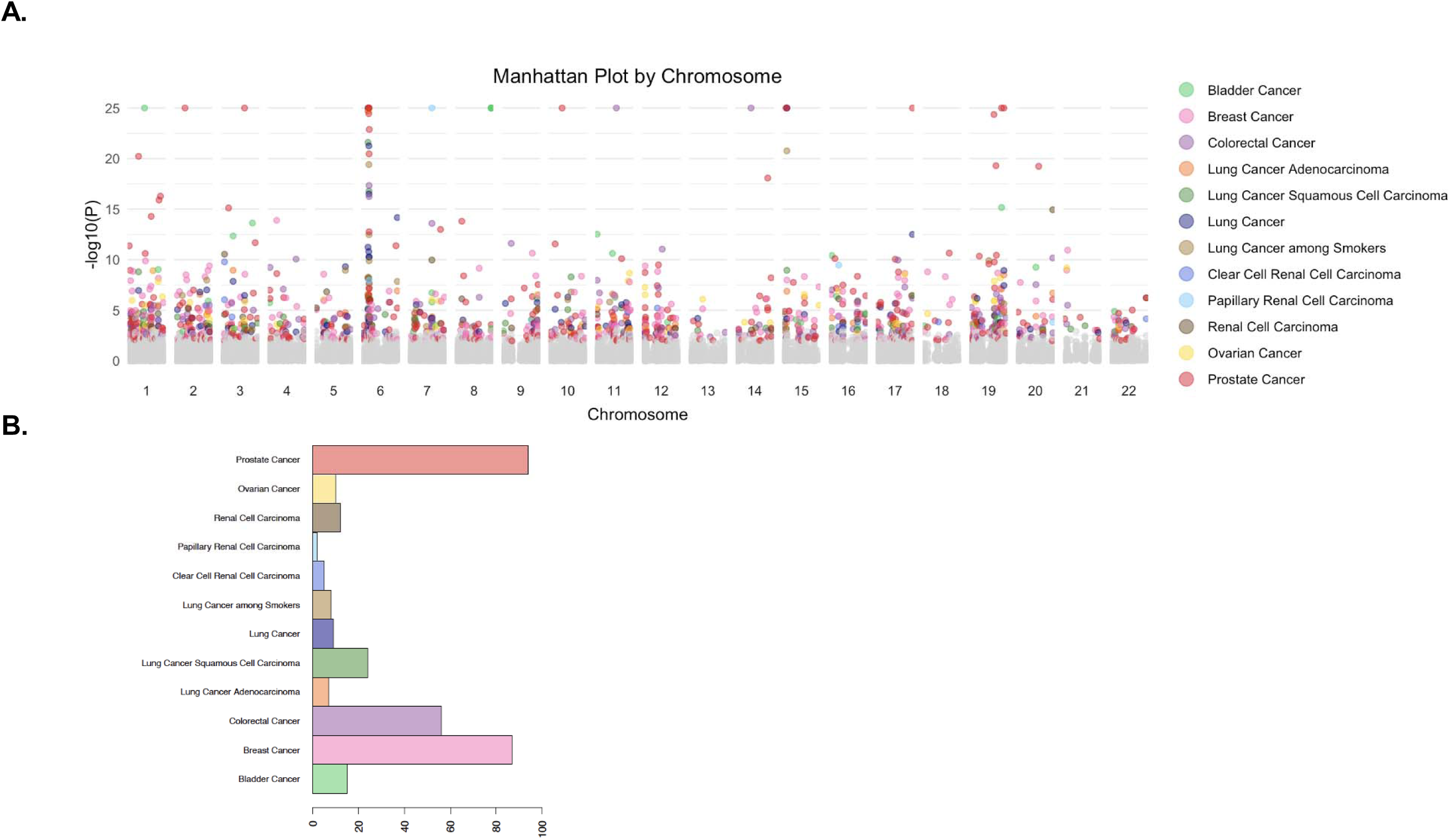
Effect of PA-associated proteins on cancers. (A) Manhattan plot for the associations of PA-associated proteins with different cancers colored by the respective cancers. Associations with p-values < 1×10^−25^ was trimmed to 1×10^−25^ for the ease of viewing. (B) Barplot showing the number of unique proteins associated to each cancer at FDR < 0.05.

Consistent with habitual PA acting through broadly shared as well as tissue-specific biology, 74 proteins were associated with more than one cancer or subtype. Of these, B2M and CD74, both reported as Tier-1 cancer genes in COSMIC^36^, were associated with 6 cancers or subtypes, while 5 proteins (BTN3A2, MUC2, BTN2A1, IFNL1 and SELE) were associated with 5 cancers or subtypes. Additionally, several of these proteins had opposing effects on different cancers suggesting context dependent effects. For example, MUC2, inversely impacted by total log acceleration activity (TLAC_mean; β=-0.09, p=2.2×10^−3^) is also inversely associated with genitourinary cancers (bladder and kidney) while positively associated with breast, prostate and colorectal cancers (**Table S6**).

To illustrate how the two-stage framework prioritizes cancer-relevant, PA-responsive proteins, we examine prostate cancer and renal cell carcinoma in detail.

### Prostate cancer (PCa)

Of the 213 candidate intermediates, 94 were associated with PCa risk (**Figure 4A**; **Table S6**), the largest set among the cancers examined. Among these, several have established roles in prostate biology and PCa. This includes, Microseminoprotein Beta (MSMB), a protein secreted by the prostate gland^37,38^ which was associated with lower risk for PCa (β = −0.21, p < 10^−125^), consistent with its reported protective role. In stage 1, we found that longer duration of light-intensity activity was associated with higher MSMB levels (β=0.13, p=9.48×10^−5^) describing a coherent chain in which greater habitual light activity corresponds to higher MSMB and, in turn, lower PCa risk. We also identified a positive association between genetically predicted TNF levels with PCa risk (β=0.13, p=3.3×10^−05^)^39,40^, while total activity volume was inversely associated with TNF (TLAC_mean; β=-0.13, p=3.6×10^−04^) from stage 1. Higher sedentary time was associated with higher TNF (LST_mean; β=0.07, p=2.2×10^−03^), opposite in direction to activity and consistent with a coherent pattern in which greater habitual activity, and less sedentary time, lowers TNF and thereby PCa risk.

**Figure 4:**
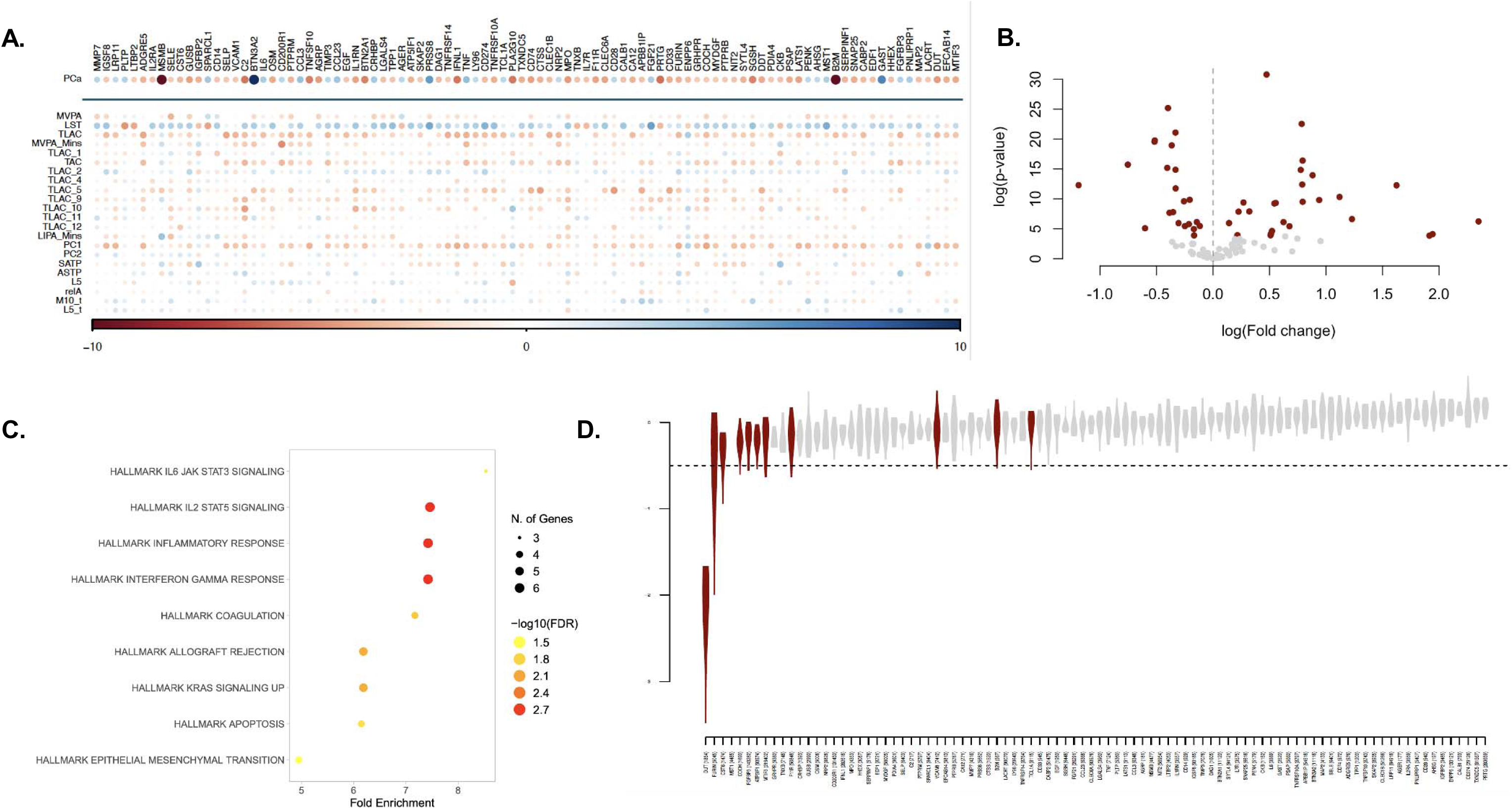
Association of PA-associated proteins with risk of Prostate cancer (PCa). Heatmap showing the Z-values of MR association between genetically predicted 94 PA-associated proteins with significant effect on PCa, across 22 different measures of PA an PCa. The size and intensity of colors in bubbles are proportional to the strength of association with negative values in red and positive in blue. (B) Differential expression of the genes corresponding to the 94 proteins in TCGA-PRAD. (C) Top enriched pathways for the 94 prot ins among Hallmark pathways (D) Violin plots of CRISPR essentiality scores (CERES) across 9 PCa cell lines with the proteins having at least one score < −0.5 marked in red.

The 94 PCa-associated proteins were enriched for cancer relevance across independent resources. Nine (9.6%) were Tier-1 cancer genes in COSMIC and 21 (22.3%) were candidate cancer driver genes in the Network of Cancer Genes^36,41^ and 48 (51.1%) exhibited differential expression compared to matched normal tissues in The Cancer Genome Atlas (TCGA)-prostate adenocarcinoma (PRAD; **Figure 4B; Table S7**)^42^. Taken together, the 94 proteins are enriched in several known cancer related pathways (**Figure 4C**) including PI3K/AKT signalling (FDR=1.7×10^−3^), KRAS signalling (FDR = 9.0×10^−3^) and apoptosis (FDR = 2.0×10^−2^). Additionally, 11 genes of the corresponding proteins were identified to be essential for cell proliferation in at least CRISPR knockout experiments^43^ in prostate cancer cell lines, indicating their relevance to PCa (**Figure 4D**).

### Renal cell carcinoma (RCC)

Twelve candidate intermediates were associated with renal cell carcinoma (RCC) risk (**Figure 5A; Table S6**); seven were associated with at least one accelerometer-derived activity measure and five with self-reported sedentary time. Unlike prostate cancer, the implied direction of the PA effect was predominantly non-protective. For most RCC proteins, the two-stage estimates were consistent with higher habitual activity or lower sedentary behavior corresponding to higher rather than lower RCC risk. This is consistent with the weak and inconsistent epidemiologic association between PA and RCC and indicates that these proteins should be interpreted as RCC-relevant, PA-responsive candidates rather than as evidence of a protective pathway.

**Figure 5:**
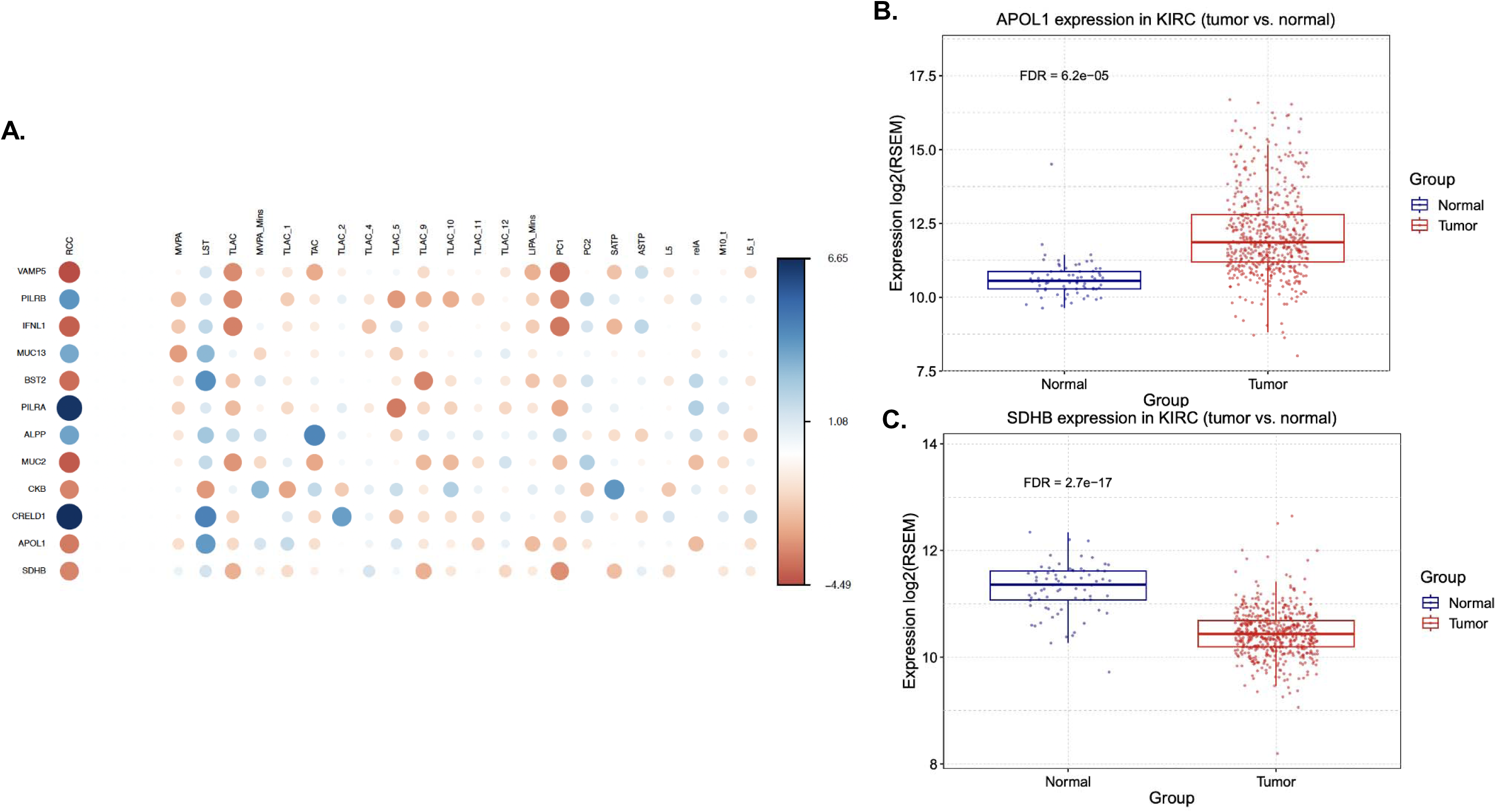
Effect of PA-associated proteins on Renal cell carcinoma (RCC). Heatmap showing the Z-values of MR association between genetically predicted levels of 12 PA associated proteins with significant effect on RCC, across 22 different measures of PA an RCC. The size and intensity of colors of bubbles are proportional to the strength of association with negative values in red and positive in blue. (B-C) Differential expression of (B) APOL1 and (C) SDHB in TCGA-KIRC with boxplots showing the gene expression values of the corresponding genes in tumor (red) and normal (blue) samples.

Apolipoprotein L1 (APOL1) which has an established link to kidney biology^44^ illustrates this pattern. APOL1 was inversely associated with RCC risk (β=-0.05, p=4.1×10^−04^), and in Stage 1, inversely associated with sedentary time (β =-0.08, p =1.9×10^−04^). Together these imply that greater sedentary behavior lowers APOL1, a protein associated with lower RCC risk which is chain consistent with the positive overall MR estimate of sedentary time on RCC (β = 0.15, p = 1.4×10^−03^). Querying TCGA-Kidney clear cell renal carcinoma gene expressions^42^, we found that APOL1 was differentially expressed in kidney tumors as well (**Figure 5B**).

Succinate dehydrogenase subunit B (SDHB), whose germline mutations cause a hereditary subtype of RCC^45,46^, and which is a Tier-1 cancer gene in COSMIC was inversely associated with RCC risk (β=-0.16, p=7.7×10^−04^), and positively with PC1, the first principal component of the log-transformed minute-level activity profiles (β=0.12, p=1.7×10^−03^). Because the overall PC1_mean–RCC association was null (β = −0.009, p =0.9) we present SDHB as a prioritized RCC-relevant protein rather than as evidence of a PA-mediated pathway. SDHB showed lower expression in kidney tumors than in matched normal tissue in TCGA-KIRC consistent with its tumor-suppressor role. (**Figure 5C**).

Among the 12 RCC-related proteins, 9 (75%) proteins exhibited differential expression in TCGA-Kidney clear cell carcinoma (KIRC). Additionally, 11 genes of the corresponding proteins had significant correlation with immune infiltration of different cell types in kidney tumors^47,48^ indicating the immunoregulatory potential of these (**Table S8**).

## Discussion

Increased PA is consistently associated with reduced risk of several common cancers^49^, but the intermediate molecular phenotypes and mechanisms that connect PA to carcinogenesis remain incompletely characterized. Multi-omics, especially proteomics, provide a powerful approach to interrogate such intermediate processes because circulating proteins reflect integrated responses across tissues, cell types, and signaling networks and are directly relevant to both tumor biology and therapeutic targeting. In this study, we used a two-stage Mendelian randomization (TSMR) framework^14,15^ to link 22 accelerometer- and questionnaire-based PA traits to 2,940 plasma proteins and multiple cancers. We identified 503 proteins whose levels were associated with PA and, among these, 42% exhibited inferred effects on cancer risk. To our knowledge, this is the first proteome-wide, genetically anchored analysis of PA in relation to cancer, and it moves beyond overall PA-cancer associations to provide a structured, mechanistic map of PA-responsive proteins, pathways, and cell types that enhances interpretation of epidemiologic findings. By anchoring on germline instruments, our analysis prioritizes proteins most likely to reflect sustained, habitual activity rather than the transient response to acute exercise.

This distinction between habitual and acute exposure is the conceptual basis of the study. Genetic instruments for PA are fixed from conception and index a lifelong propensity toward higher activity and lower sedentary behavior, whereas exercise-intervention studies measure short-term responses to prescribed, often higher-intensity exercise. These are related but non-interchangeable exposures, and for a long-latency disease such as cancer which is primarily driven by cumulative rather than momentary influences, the habitual exposure is the more relevant one. Our benchmarking analysis supports the validity of this approach. Of 84 curated exercise-responsive proteins with genetic instruments, 72 showed nominal MR association with at least one PA measure, an enrichment robust even under the conservative assumption of fully independent measures. Because the genetic and experimental approaches rest on different designs and biases, this convergence of two distinct lines of analyses, supports the validity of PA-focused MR while not implying that genetically proxied liability is equivalent to a deliberate change in activity.

Consistent with PA being a multifaceted exposure, accelerometer-derived activity and self-reported sedentary behavior produced overlapping but distinct proteomic signatures, and sedentary behavior was not simply the inverse of activity. The implicated proteins were enriched for inflammatory, metabolic, immune, and vascular pathways and were differentially expressed following exercise in transcriptomic data, predominantly in muscle and across immune cell populations, situating them in biology plausibly relevant to carcinogenesis. Therefore, linking these proteins to cancer illustrated the value. For prostate cancer, MSMB and TNF provided coherent chains from habitual activity to risk, but across the 94 prostate-associated proteins the implied direction of the PA effect was evenly split between risk-lowering and risk-raising; for RCC, the majority of proteins, including APOL1, implied a non-protective direction. These patterns are consistent with the weak and inconsistent epidemiologic associations of PA with these two cancers, and they underscore that the prioritized proteins represent testable, context-specific hypotheses rather than uniform evidence that activity lowers risk at any given site. Multi-cancer proteins such as B2M, CD74, and MUC2, the last with opposing directions across sites, further point to shared but tissue-dependent pathways.

Several limitations have direct consequences on interpretation. First, MR estimates for a behavioral exposure are difficult to interpret causally. Genetic instruments for PA are individually weak, act from early life, and may influence activity through correlated behavioral, metabolic, and psychological pathways, so the gene-environment equivalence assumption cannot be taken for granted, and associations may reflect broader correlates of activity rather than activity itself. Second, forward MR cannot fully exclude reverse or bidirectional effects or shared upstream determinants at either stage. Third, horizontal pleiotropy is likely widespread for a behavioral trait. HEIDI outlier removal and MR-Egger reduce but do not eliminate this, and robustness to pervasive pleiotropy, to stricter instrument and LD thresholds, and formal mediation and multivariable analyses remain important next steps. Fourth, the two-stage design screens rather than formally partitions mediation: proteins supported in both stages are candidate intermediates, and because the same protein GWAS informs both stages, Stage 2 results are best read as prioritization within a pre-selected set rather than independently calibrated tests. Finally, analyses were largely restricted to European-ancestry GWAS, the Olink platform captures a targeted subset of the proteome, and aggregated cancer endpoints may obscure subtype heterogeneity.

Despite these caveats, our work underscores the interpretive gains of embedding PA–cancer epidemiology within a proteogenetic framework. By systematically linking different facets of PA to plasma proteins, cell types, pathways, and cancer outcomes, we move beyond simplistic understanding of PA on cancers, to a more nuanced understanding of which biological programs are altered by PA and how they may translate into reduced cancer risk. This framework is explicitly hypothesis generating. Our analysis attempts to shorten the search space from proteome-wide scale to a tractable set of potentially PA-associated candidates with genetic and functional support that merit mechanistic experiments, validation in intervention studies, and evaluation as potential biomarkers or therapeutic targets. This study provides a prioritized catalog of PA-responsive proteins and pathways that can guide experimental follow-up, inform the design and monitoring of PA interventions, and ultimately help to refine prevention strategies that leverage exercise as a modifiable, low-cost tool in cancer control.

## Data Availability

GWAS summary statistics for PA and cancers are available on GWAS catalog (https://www.ebi.ac.uk/gwas/) and those for plasma proteins are available at (https://www.synapse.org/Synapse:syn51364943). GSCA for downstream analysis is available online at https://guolab.wchscu.cn/GSCA/ including TCGA differential expression and immune infiltration data. CRISPR cell essentiality data is available at DepMap: https://depmap.org/portal/. Pathway analysis was conducted using ShinyGO: https://bioinformatics.sdstate.edu/go/. ExerGeneDB database can be accessed at: https://exergenedb.com/

## Acknowledgements

The contributions of the NIH author(s) are considered Works of the United States Government. The findings and conclusions presented in this paper are those of the author(s) and do not necessarily reflect the views of the NIH or the U.S. Department of Health and Human Services.

## Code availability

GSMR v1.94.1: https://yanglab.westlake.edu.cn/software/gcta/ MRlap v0.0.3: https://github.com/n-mounier/MRlap MendelianRandomization v0.10.0: 10.32614/CRAN.package.MendelianRandomization TwoSampleMR v0.6.21: https://mrcieu.github.io/TwoSampleMR/index.html

## References

1. Sud, A., Kinnersley, B. & Houlston, R. S. Genome-wide association studies of cancer: current insights and future perspectives. Nat. Rev. Cancer 17, 692–704 (2017).

2. GBD 2019 Cancer Risk Factors Collaborators. The global burden of cancer attributable to risk factors, 2010-19: a systematic analysis for the Global Burden of Disease Study 2019. Lancet Lond. Engl. 400, 563–591 (2022).

3. Arani, G. et al. Plasma Proteomic Signatures of Physical Activity Provide Insights into Biological Impacts of Physical Activity and its Protective Role Against Dementia. MedRxiv Prepr. Serv. Health Sci. 2025.01.16.25320290 (2025) doi:10.1101/2025.01.16.25320290.

4. Chen, X. et al. Causal relationship between physical activity, leisure sedentary behaviors and COVID-19 risk: a Mendelian randomization study. J. Transl. Med. 20, 216 (2022).

5. Park, S. et al. Causal effects of physical activity or sedentary behaviors on kidney function: an integrated population-scale observational analysis and Mendelian randomization study. Nephrol. Dial. Transplant. Off. Publ. Eur. Dial. Transpl. Assoc. - Eur. Ren. Assoc. 37, 1059–1068 (2022).

6. Warburton, D. E. R., Nicol, C. W. & Bredin, S. S. D. Health benefits of physical activity: the evidence. CMAJ Can. Med. Assoc. J. J. Assoc. Medicale Can. 174, 801–809 (2006).

7. Courneya, K. S. et al. Structured Exercise after Adjuvant Chemotherapy for Colon Cancer. N. Engl. J. Med. 393, 13–25 (2025).

8. Sun, B. B. et al. Plasma proteomic associations with genetics and health in the UK Biobank. Nature 622, (2023).

9. Zhang, J. et al. Plasma proteome analyses in individuals of European and African ancestry identify cis-pQTLs and models for proteome-wide association studies. Nat. Genet. 54, (2022).

10. Gadd, D. A. et al. Blood protein assessment of leading incident diseases and mortality in the UK Biobank. Nat. Aging 4, 939–948 (2024).

11. Zhao, J. H. et al. Genetics of circulating inflammatory proteins identifies drivers of immune-mediated disease risk and therapeutic targets. Nat. Immunol. 24, 1540–1551 (2023).

12. Western, D. et al. Proteogenomic analysis of human cerebrospinal fluid identifies neurologically relevant regulation and implicates causal proteins for Alzheimer’s disease. Nat. Genet. 56, 2672–2684 (2024).

13. Zhou, R. et al. Genetic associations of circulating plasma proteins with cardiometabolic diseases. Genome Biol. 10.1186/s13059-026-03962-x (2026) doi:10.1186/s13059-026-03962-x.

14. Yoshiji, S. et al. Proteome-wide Mendelian randomization implicates nephronectin as an actionable mediator of the effect of obesity on COVID-19 severity. Nat. Metab. 5, 248–264 (2023).

15. Yoshiji, S. et al. Integrative proteogenomic analysis identifies COL6A3-derived endotrophin as a mediator of the effect of obesity on coronary artery disease. Nat. Genet. 57, 345–357 (2025).

16. Smith, G. D. & Ebrahim, S. ‘Mendelian randomization’: Can genetic epidemiology contribute to understanding environmental determinants of disease? Int. J. Epidemiol. 32, (2003).

17. Burgess, S., Butterworth, A. & Thompson, S. G. Mendelian randomization analysis with multiple genetic variants using summarized data. Genet. Epidemiol. 10.1002/gepi.21758 (2013) doi:10.1002/gepi.21758.

18. Burgess, S. et al. Guidelines for performing Mendelian randomization investigations: update for summer 2023. Wellcome Open Res. 4, (2023).

19. Richmond, R. C. & Davey Smith, G. Mendelian Randomization: Concepts and Scope. Cold Spring Harb. Perspect. Med. 12, a040501 (2022).

20. The 1000 Genomes Project Consortium. A global reference for human genetic variation. Nature 526(7571), 68–74 (2015).

21. Zhu, Z. et al. Causal associations between risk factors and common diseases inferred from GWAS summary data. Nat. Commun. 9, (2018).

22. Qi, G. et al. Genome-wide association studies of 27 accelerometry-derived physical activity measurements identified novel loci and genetic mechanisms. Genet. Epidemiol. 46, (2022).

23. Wang, Z. et al. Genome-wide association analyses of physical activity and sedentary behavior provide insights into underlying mechanisms and roles in disease prevention. Nat. Genet. 54, 1332–1344 (2022).

24. Yengo, L. et al. Meta-analysis of genome-wide association studies for height and body mass index in ∼700 000 individuals of European ancestry. Hum. Mol. Genet. 27, (2018).

25. Bycroft, C. et al. The UK Biobank resource with deep phenotyping and genomic data. Nature 562, 203–209 (2018).

26. Mounier, N. & Kutalik, Z. Bias correction for inverse variance weighting Mendelian randomization. Genet. Epidemiol. 47, 314–331 (2023).

27. Robbins, J. M. et al. Human plasma proteomic profiles indicative of cardiorespiratory fitness. Nat. Metab. 3, 786–797 (2021).

28. Wei, W., Raun, S. H. & Long, J. Z. Molecular Insights From Multiomics Studies of Physical Activity. Diabetes 73, 162–168 (2024).

29. Guseh, J. S. et al. An expanded repertoire of intensity-dependent exercise-responsive plasma proteins tied to loci of human disease risk. Sci. Rep. 10, 10831 (2020).

30. Kuramoto, K., Liang, H., Hong, J.-H. & He, C. Exercise-activated hepatic autophagy via the FN1-α5β1 integrin pathway drives metabolic benefits of exercise. Cell Metab. 35, 620–632.e5 (2023).

31. Wei, W. et al. Organism-wide, cell-type-specific secretome mapping of exercise training in mice. Cell Metab. 35, 1261–1279.e11 (2023).

32. Houseknecht, K. L., Baile, C. A., Matteri, R. L. & Spurlock, M. E. The biology of leptin: a review. J. Anim. Sci. 76, 1405–1420 (1998).

33. Qian, H. et al. Uncovering the Molecular Landscape of Physical Activity: Proteomic Insights from the UK Biobank. MedRxiv Prepr. Serv. Health Sci. 2025.06.27.25330418 (2025) doi:10.1101/2025.06.27.25330418.

34. Burgess, S., Dudbridge, F. & Thompson, S. G. Combining information on multiple instrumental variables in Mendelian randomization: Comparison of allele score and summarized data methods. Stat. Med. 35, (2016).

35. Pan, L. et al. ExerGeneDB: A physical exercise-regulated differential gene expression database. J. Sport Health Sci. 14, 101027 (2025).

36. Sondka, Z. et al. The COSMIC Cancer Gene Census: describing genetic dysfunction across all human cancers. Nat. Rev. Cancer 18, (2018).

37. Abrahamsson, P. A., Lilja, H., Falkmer, S. & Wadström, L. B. Immunohistochemical distribution of the three predominant secretory proteins in the parenchyma of hyperplastic and neoplastic prostate glands. The Prostate 12, 39–46 (1988).

38. Sjöblom, L. et al. Microseminoprotein-Beta Expression in Different Stages of Prostate Cancer. PloS One 11, e0150241 (2016).

39. Wang, X. & Lin, Y. Tumor necrosis factor and cancer, buddies or foes? Acta Pharmacol. Sin. 29, 1275–1288 (2008).

40. Balkwill, F. Tumour necrosis factor and cancer. Nat. Rev. Cancer 9, 361–371 (2009).

41. Repana, D. et al. The Network of Cancer Genes (NCG): A comprehensive catalogue of known and candidate cancer genes from cancer sequencing screens. Genome Biol. 20, (2019).

42. Weinstein, J. N. et al. The cancer genome atlas pan-cancer analysis project. Nat. Genet. 45, (2013).

43. Tsherniak, A. et al. Defining a Cancer Dependency Map. Cell 170, 564–576.e16 (2017).

44. Hu, C.-A. A., Klopfer, E. I. & Ray, P. E. Human apolipoprotein L1 (ApoL1) in cancer and chronic kidney disease. FEBS Lett. 586, 947–955 (2012).

45. Williamson, S. R. et al. Succinate dehydrogenase-deficient renal cell carcinoma: detailed characterization of 11 tumors defining a unique subtype of renal cell carcinoma. Mod. Pathol. Off. J. U. S. Can. Acad. Pathol. Inc 28, 80–94 (2015).

46. Ricketts, C. J. et al. Succinate dehydrogenase kidney cancer: an aggressive example of the Warburg effect in cancer. J. Urol. 188, 2063–2071 (2012).

47. Liu, C.-J. et al. GSCA: an integrated platform for gene set cancer analysis at genomic, pharmacogenomic and immunogenomic levels. Brief. Bioinform. 24, bbac558 (2023).

48. Miao, Y. R. et al. ImmuCellAI: A Unique Method for Comprehensive T-Cell Subsets Abundance Prediction and its Application in Cancer Immunotherapy. Adv. Sci. 7, (2020).

49. McTiernan, A. et al. Physical Activity in Cancer Prevention and Survival: A Systematic Review. Med. Sci. Sports Exerc. 51, 1252–1261 (2019).

