## Supplementary Methods for "Proteogenomic analysis provides insights into the effect of physical activity on cancers"

**Protein-protein enrichment (PPI)**. PPI was conducted using data from STRINGdb to identify whether PA-related proteins exhibited more interactions than expected by chance, with significance indicating functional connectivity within the protein set (See **Data Availability**).

**Pathway enrichment**. Pathway enrichment was used to identify which gene-set/biological processes had significant overrepresentation of (A) PA-related proteins and (B) proteins related to PCa. Using multiple pathway databases including Hallmark genes, KEGG, Gene ontology and others, curated in MSigDB, we used ShinyGO online portal to identify pathways enriched for a query set of genes/proteins.

**Differential expression in response to exercise.** ExerGeneDB curates a list of experiments in humans and mice, listing genes that exhibit altered expression in response to an exercise/PA stimulus. The p-value for differential expression was corrected using Benjamini-Hochberg false discovery rate correction to obtain p_adj_. Genes with p_adj_ < 0.05 were reported and highlighted.

**Differential expression for cancers**. Differential expression analyses was conducted to identify genes with significantly altered expression levels between matched tumor and normal samples in a given cancer. The p-value was then corrected using Benjamini-Hochberg false discovery rate correction to obtain p_adj_. Genes with p_adj_ < 0.05 were reported and highlighted.

**Cancer genes**. We curated a list of canonical or candidate cancer driver genes from the Network of Cancer Genes & Healthy Drivers v7.0 as well as a set of cancer driver genes as reported in COSMIC. We queried this list to identify which PA-related proteins were also reported to cancer driver or candidate cancer driver genes.

**CRISPR essentiality**. To investigate the effect of the cognate genes of proteins on essentiality for proliferation and survival of cancer cells, we used the Achilles dataset from the DepMap portal (See **Data Availability**). This provides estimated gene-dependency levels from CRISPR–Cas9 essentiality screens for a total 18,119 genes using a computational method, CERES. In a given cell line, relevant to for the cancer being analyzed, we identified the cognate genes for a query list of proteins that had CERES value of −0.5 or lower as recommended, indicating essentiality for cell proliferation. This was done for prostate cancer cell lines and renal cell carcinoma cell lines.

**Supplementary Figure**: Comparison of effect size estimates obtained from GSMR and effect size estimates from MRlap correcting for bias due to sample overlap. Associations with significant differences (p_diff_ < 0.005) are colored in red.

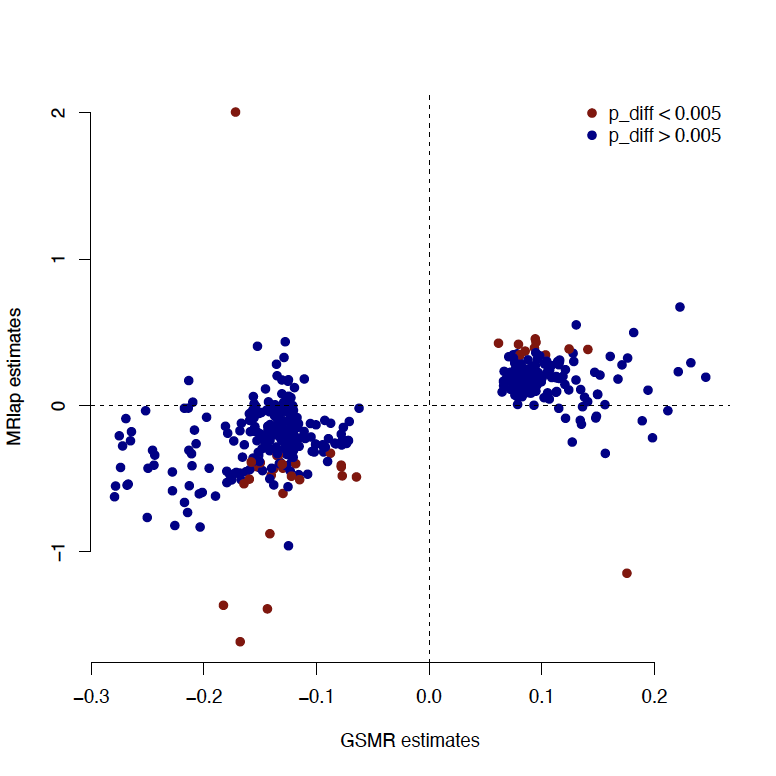
